# Menstrual Cycle Changes among Reproduction-Aged Iranian Women Following COVID-19 Vaccination

**DOI:** 10.64898/2026.07.07.26357499

**Authors:** Afrooz Azad, Fatemeh Darsareh, Mahbobe Ebrahimi abshur, Mojgan Hajisafari, Azadeh Mahmoudi Essaabadi

## Abstract

**Background:** Menstrual cycle disturbances have been increasingly reported after COVID-19 vaccination, raising questions about their prevalence and clinical significance among women of reproductive age.

**Objective:** This study aimed to investigate the incidence and types of menstrual cycle alterations following different doses of COVID-19 vaccines among Iranian women of reproductive age.

**Methods:** A cross-sectional survey was conducted among vaccinated women who reported their menstrual cycle status before and after each vaccine dose. Data on cycle regularity, flow characteristics, and specific menstrual disorders were collected and analyzed.

**Results:** Menstrual cycle alterations were reported by 28.8%, 25.4%, 30.3%, and 68.4% of participants after the first, second, third, and fourth vaccine doses, respectively. The most common changes were oligomenorrhea after the first and second doses (8.9% and 5.6%), menorrhagia after the third dose (5.3%), and hypomenorrhea after the fourth dose (8.3%). Comparisons with international studies revealed a wide variation in prevalence (ranging from 25% to 78%), which may be explained by differences in methodology, population characteristics, vaccine types, and pre-vaccination health status.

**Conclusion:** A considerable proportion of Iranian women experienced menstrual alterations following COVID-19 vaccination, most commonly oligomenorrhea, menorrhagia, and hypomenorrhea. While generally self-limiting, these findings highlight the need to integrate menstrual health into post-vaccination monitoring and patient counseling. Future research should explore the underlying immune–endocrine mechanisms and long-term clinical implications of these changes.

## Introduction

The COVID-19 pandemic has significantly influenced many areas of human life in recent years(1). Vaccination has emerged as the most effective strategy against SARS-CoV-2. Nonetheless, research has documented a variety of vaccine-associated side effects, ranging from mild reactions such as fever, headache, fatigue, and localized arm pain to more serious complications, including stroke, thrombosis, and anaphylaxis(2).

The female menstrual cycle is precisely orchestrated by the hypothalamic–pituitary–ovarian (HPO) axis. Perturbations within this regulatory system may occur secondary to disturbances in physiological homeostasis, which can be induced by infectious and febrile conditions, psychological stressors, or the administration of particular pharmacological agents(3). Following the emergence of COVID-19, numerous women, regardless of whether they had been vaccinated, reported disruptions in their menstrual cycles. According to a statement released in August 2021 by the Pharmacovigilance Risk Assessment Committee (PRAC) of the European Medicines Agency (EMA), no evidence was identified to suggest a link between vaccination and menstrual cycle irregularities(4). The Medicines and Healthcare products Regulatory Agency (MHRA), drawing on data from more than 41,900 women, documented instances of unusually heavy menstrual bleeding, alterations in menstrual cycles, or unanticipated vaginal bleeding after COVID-19 vaccination(5). Recent research indicates that SARS-CoV-2 infection, COVID-19 vaccines, and pandemic-related stress may influence menstrual cycles(6). According to one study’s findings, the prevalence of potential menstrual cycle disruption symptoms included menstrual pain (14.1%), increased menstrual frequency (9.4%), heavy menstrual bleeding (8.9%), extended menstrual duration (8.5%), amenorrhea (3.4%), exacerbation of symptoms compared to baseline (1.7%), and intermenstrual bleeding (1.1%)(1). Another study documented symptoms such as heavier-than-normal menstrual bleeding, prolonged menstruation, shorter or longer intervals between periods, intermenstrual spotting, intensified menstrual pain, menstrual pain without bleeding, and additional pelvic symptoms following COVID-19 vaccination(4).

Despite widespread concerns about a possible association between COVID-19 vaccination and menstrual cycle disturbances, clinical trial outcomes involving women aged 18 to 45 in the United States demonstrated that menstrual cycle duration remained within normal ranges(7). Prolonged menstrual irregularities may result in various complications, such as irritability and anger, depression and feelings of despair, anxiety, fatigue, alterations in appetite and sleep patterns, and joint and muscle pain(8). The study included 170 women from two cities of Iran, equally divided between 85 participants from Hamedan and 85 from Zahedan. Findings revealed that approximately 57.6% of women in Hamedan and 54.1% in Zahedan experienced menstrual disorder symptoms after receiving the COVID-19 vaccine. Moreover, disruptions in menstrual cycle duration were more prevalent in Hamedan. Additionally, irregularities in the severity and recurrence of menstruation were more commonly reported in Hamedan than in Zahedan(9). These results enhance our understanding of the regional and demographic influences on menstrual alterations following vaccination. Given the demographic diversity in Iran and to enhance the available evidence, this study was formulated to explore the prevalence of menstrual cycle alterations and their associated symptoms among women in Iran.

## Methods

### Study Design

This investigation was structured as a cross-sectional descriptive study aimed at evaluating the prevalence of menstrual cycle alterations and related symptoms among Iranian women aged 18 to 45 years who had received at least one dose of a COVID-19 vaccine. The study was initiated after securing ethical approval from the Ethics Committee of Hormozgan University of Medical Sciences and obtaining all requisite permissions.

### Study Population and Sampling

The study population consisted of all women aged 18 to 45 years in Iran who had received at least one dose of a COVID-19 vaccine. Sample size calculation was based on the proportion estimation formula, referencing a 52% prevalence of menstrual irregularities reported by Nazir et al.(10). With a type I error (α) of 0.05, 80% power, and a precision (d) of 0.04, a sample size of 986 participants was determined using MedCalc version 14.

A convenience sampling approach was employed, whereby eligible women aged 18 to 45 years who had received at least one vaccine dose and provided consent to participate were recruited until the target sample size was achieved.

### Inclusion Criteria

- Age ranging from 18 to 45 years
- Administration of at least one COVID-19 vaccine dose
- Voluntary provision of informed consent

### Exclusion Criteria

- Pregnancy
- Breastfeeding
- Prior hysterectomy
- Use of hormonal contraceptives
- BMI below 18 or above 30 kg/m²
- Diagnosed thyroid or prolactin hormonal disorders

### Data Collection

A custom-designed questionnaire was developed to collect relevant data, covering:

- Demographic details: age, height, weight, employment status, educational background, and residential location
- Vaccination history: number of COVID-19 vaccine doses and type of vaccine administered per dose
- Menstrual cycle information: nature and types of menstrual cycle changes post-vaccination, and the time elapsed between vaccination and the onset of these changes

The questionnaire was hosted on the Prosline platform, with its access link distributed via social media platforms such as WhatsApp, Telegram, Eitaa, and others. It remained accessible for responses for over 8 weeks. Following data collection, responses were exported to SPSS version 26 for statistical processing.

### Data Analysis

Descriptive statistics, including means, percentages, and frequency distributions, were employed to characterize the study population and estimate the prevalence of menstrual cycle changes. Prevalence confidence intervals were computed using the pii command in Stata version 14. Associations between menstrual cycle alterations and variables such as age, occupation, residence, and vaccine type were assessed using chi-square tests when applicable, or Fisher’s exact tests when assumptions for chi-square were not met. A P-value threshold of <0.05 was used to determine statistical significance. Visual representations of the data were created using Microsoft Excel.

## Ethical Considerations

The study adhered to ethical guidelines, with approval obtained from the Ethics Committee of Hormozgan University of Medical Sciences(ethical codeIR.HUMS.REC.1402.395). Informed consent was secured from all participants, and measures were implemented to ensure data confidentiality and participant anonymity throughout the research process.

## Results

### Demographic Characteristics

The study included 986 women aged 18 to 45 years, with a mean age of 31.72 ± 7.57 years. The demographic characteristics of the participants are summarized in Table 1.

**Table 1:** Demographic Characteristics of Study Participants.

| **Variable** | **Description** | **Value** |
| --- | --- | --- |
| **Age (years)** | Mean ± SD | $31.72 \pm 7.57$ |
| **Height (cm)** | Mean ± SD | $161.61 \pm 7.71$ |
| **Weight (kg)** | Mean ± SD | $65.04 \pm 10.22$ |
| **BMI (kg/m<sup>2</sup>)** | Mean ± SD | $24.86 \pm 3.19$ |
| **Place of Residence** | Urban | 743 (75.4%) |
|  | Rural | 243 (24.6%) |
| **Employment Status** | Unemployed/Housewife | 445 (45.1%) |
|  | Employed | 541 (54.9%) |
| **Education Level** | Illiterate | 40 (4.1%) |
|  | Primary Education | 62 (6.3%) |
|  | Secondary Education | 100 (10.1%) |
|  | High School Diploma | 276 (28.0%) |
|  | University Degree | 508 (51.5%) |
| **Number of COVID-19 Vaccine Doses** | 1 Dose | 188 (19.1%) |
|  | 2 Doses | 366 (37.1%) |
|  | 3 Doses | 299 (30.3%) |
|  | 4 Doses | 133 (13.5%) |

### Menstrual Cycle Characteristics

The menstrual cycle status before the first, second, third, and fourth doses of the COVID-19 vaccine, the prevalence of menstrual cycle changes post-vaccination, and the time interval from each dose to the onset of menstrual changes were assessed. Of the 986 participants, 284 reported changes after the first dose; of the 798 participants who received at least two doses, 203 reported changes after the second dose; of the 432 participants who received at least three doses, 131 reported changes after the third dose; and of the 133 participants who received four doses, 91 reported changes after the fourth dose. Overall, 322 participants reported menstrual cycle changes after at least one dose. These findings are summarized in Table 2.

**Table 2:**
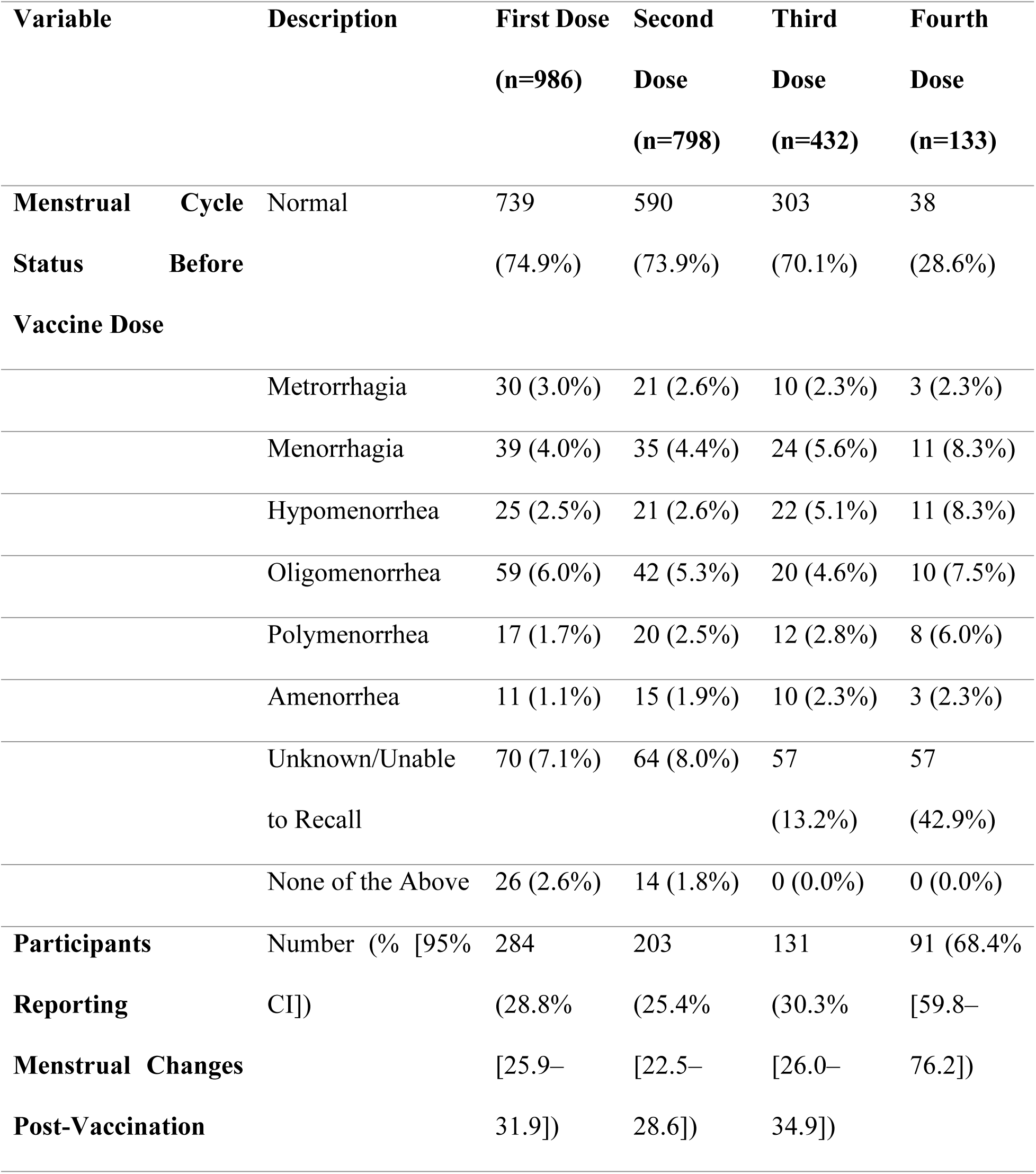
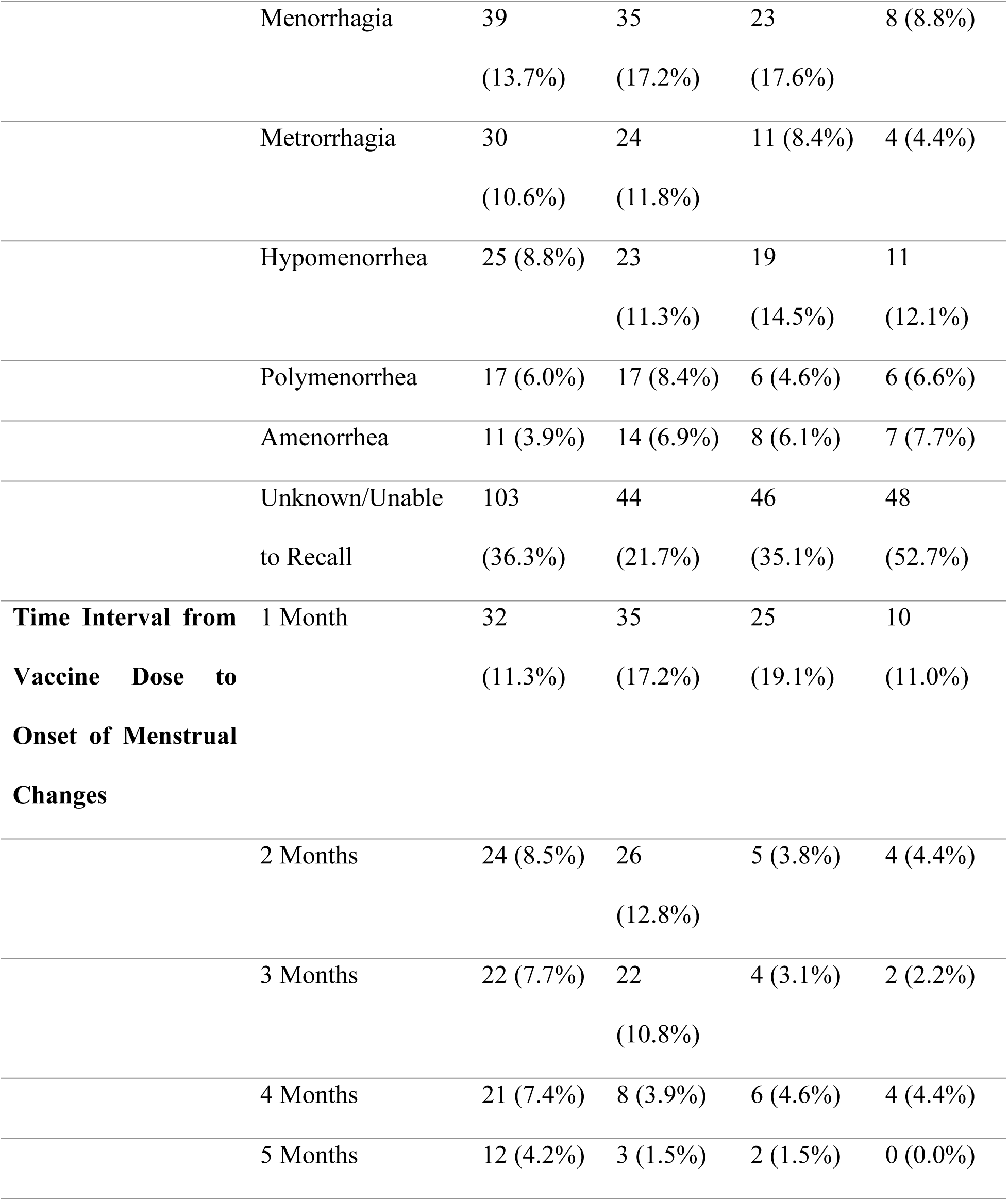
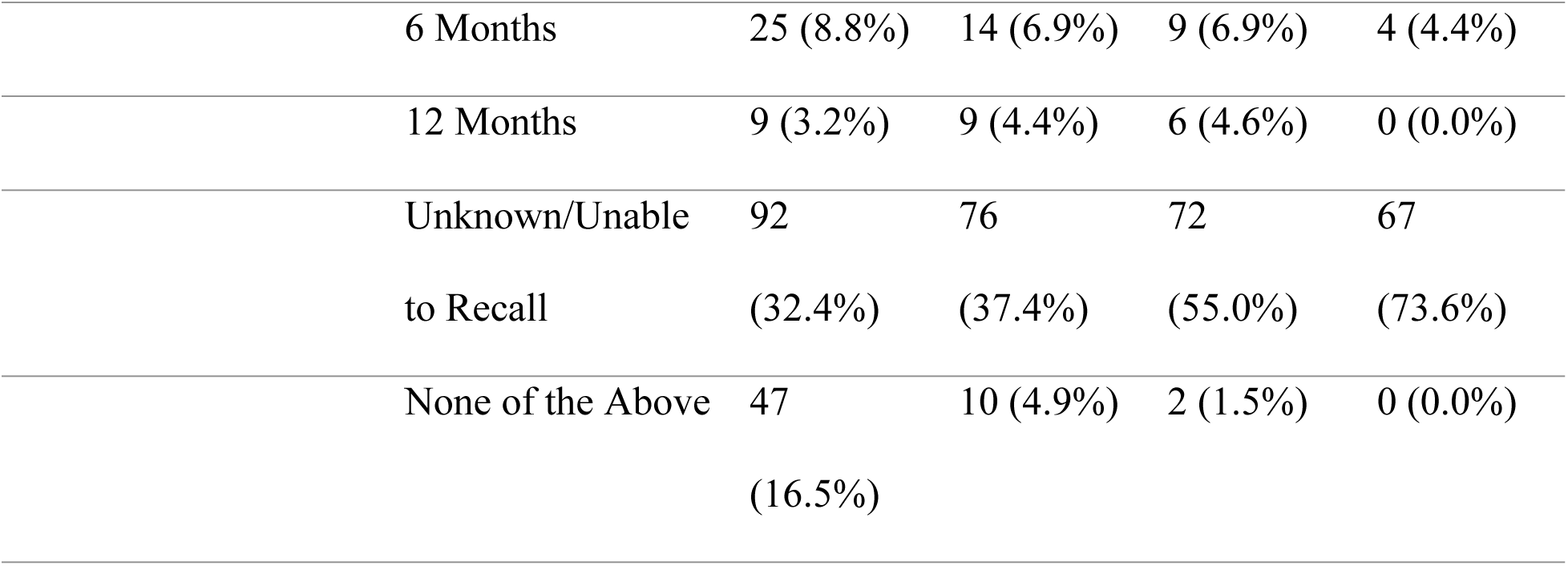
Menstrual Cycle Characteristics and Post-Vaccination Changes.

### Overall Prevalence of Menstrual Changes

Overall, 322 participants (32.7% [95% CI: 29.7–35.7]) reported menstrual cycle changes after receiving at least one dose of the COVID-19 vaccine.

A notable correlation was identified between the number of vaccine doses received and alterations in menstrual cycles, with 29.5% of those reporting menstrual changes and just 5.7% of those without such changes having completed four doses of the COVID-19 vaccine (P < 0.001).

## Discussion

The findings of this study indicated that the incidence of menstrual cycle alterations after the first, second, third, and fourth doses of the COVID-19 vaccine among the women involved was 28.8%, 25.4%, 30.3%, and 68.4%, respectively. This suggests that between one-quarter and two-thirds of the participants underwent menstrual cycle changes post-vaccination.

Previous research on the prevalence of menstrual cycle changes following COVID-19 vaccination has yielded diverse results, potentially attributable to differences in sample sizes, the approach to evaluating menstrual irregularities, the specific disorders under consideration, demographic profiles, the vaccine types used, and the pre-vaccination health status of participants. In their systematic review, Nazir and colleagues analyzed 14 studies and found that 52% of the 78,138 women examined experienced menstrual disruptions post-vaccination(10). Abdullah et al. reported that 71.8% of participants exhibited menstrual changes, especially following the second dose(11). Fallatah et al. documented menstrual cycle alterations in 54.7% of women after vaccination. Aljehani et al. observed that 41.7% (223 women) experienced changes after the first dose, increasing to 44.1% (236 women) after the second dose(12). The prevalence was 67% according to Filfilan et al.(13), 53.1% in the study by Toktas et al. from Turkey(14), 78% by Baena-Garcia et al. from Spain(15), 50–60% after the first dose and 60–70% after the second dose as per Lagana et al. from Italy (16), and 37.8% as reported by Trogstad et al. from Norway(4). Additionally, Farland et al. from the United States found that about 25% of vaccinated individuals noted menstrual cycle changes, predominantly after the second dose (56%), with 18% and 14% reported after the first and third doses, respectively(17). An important consideration is that participants in this study received vaccine doses at different intervals, making it uncertain in cases with a long latency period (e.g., six months or one year) whether reported changes after later doses were carryovers from earlier doses or new developments. Nonetheless, the menstrual cycle status of each participant was assessed before each dose. The considerable heterogeneity in the reported prevalence across studies may be attributed not only to methodological differences, but also to cultural variations in reporting menstrual symptoms, genetic predispositions, stress related to the pandemic and vaccination campaigns, and variations in vaccine brands administered across countries. Such factors highlight the importance of contextualizing findings within local populations when interpreting menstrual health outcomes.

Another key observation from this study was that the predominant menstrual cycle alterations after the first, second, third, and fourth doses of the COVID-19 vaccine were oligomenorrhea (8.9%), oligomenorrhea (5.6%), menorrhagia (5.3%), and hypomenorrhea (8.3%), respectively. Consequently, oligomenorrhea, menorrhagia, and hypomenorrhea emerged as the leading menstrual cycle variations in this research. In a similar vein, Al Kadri et al. in their meta-analysis of 16 cross-sectional studies identified menorrhagia (24.24% [95% CI: 12.8–35.6%]), oligomenorrhea (22.7% [95% CI: 13.5–32%]), and polymenorrhea (16.2% [95% CI: 10.7–21.6%]) as the most frequent menstrual irregularities(18). In contrast, Duijster et al. found that amenorrhea/oligomenorrhea was the most commonly reported menstrual effect, accounting for 33% of 24,090 cases(19). Oligomenorrhea, defined as intervals between periods exceeding 37 days, was notably the most prevalent disorder in this study, particularly following the first and second vaccine doses.

Taken together, these findings strengthen the emerging evidence that COVID-19 vaccination is associated with subtle but measurable alterations in menstrual cycle characteristics, including both bleeding patterns and cycle length. Alterations in menstrual cycle duration post-COVID-19 vaccination represent a widely studied outcome across research. Most investigations have noted significant differences in cycle length changes between vaccinated and unvaccinated groups. In an international cohort study by Edelman and colleagues, which included 19,622 participants (14,936 vaccinated), vaccinated individuals exhibited an approximate one-day increase in cycle length compared to unvaccinated individuals after both doses(20). Similarly, Gibson et al in their large cohort study of 9,652 participants (8,485 vaccinated) reported a cycle length increase of 0.50 days (95% CI: 0.22–0.78) after the first dose and 0.39 days (95% CI: 0.11–0.67) after the second dose(21). These outcomes align with the current study’s findings. Trogstad and colleagues also documented significant variations post-first and second doses(22), while Bisgaard Jensen et al. highlighted that cycle length changes were the most frequent alteration, with 9% of participants experiencing extended cycles and 7% of participants experiencing shorter cycles(23). Potential biological mechanisms underlying these alterations may involve immune–endocrine interactions triggered by vaccination. The systemic inflammatory response and subsequent cytokine release could transiently influence the hypothalamic–pituitary–ovarian (HPO) axis, thereby affecting cycle regulation. Psychological stress related to vaccination and the pandemic itself may also contribute to irregularities. Although these changes appear to be temporary in most cases, further longitudinal studies are needed to confirm their clinical significance(24, 25).

## Conclusion

This study highlights that a substantial proportion of Iranian women of reproductive age experienced menstrual cycle alterations following COVID-19 vaccination, most notably oligomenorrhea, menorrhagia, and hypomenorrhea. While such changes were generally transient and varied across vaccine doses, their high frequency underscores the importance of recognizing menstrual health as a relevant aspect of post-vaccination monitoring. Given the wide heterogeneity of findings in the global literature, cultural, biological, and methodological factors must be considered when interpreting these results. Importantly, these observations should not discourage vaccination, as the benefits of COVID-19 immunization in reducing severe disease and mortality remain unequivocal. Instead, awareness of potential menstrual changes can empower women, reduce anxiety, and guide healthcare providers in offering appropriate counseling. Future longitudinal and mechanistic studies are warranted to better understand the underlying immune– endocrine pathways and to determine the long-term clinical implications of these menstrual alterations.

## Data Availability

The data are available from the corresponding author (Azadeh Mahmoudi Essaabadi) upon reasonable request and subject to approval by the Ethics Committee of Hormozgan University of Medical Sciences. Requests for data access will be evaluated on a case-by-case basis.

## Acknowledgment

We would like to express our sincere appreciation to MD Zahra Khoshouei and MD Hossein Abdollahi from the Student Research Committee, Faculty of Medicine, Hormozgan University of Medical Sciences, Bandar Abbas, Iran for their valuable support and contributions.

## Notes

### Competing Interest Statement

The authors have declared no competing interest.

### Author Declarations

This study was approved by the Ethics Committee of Hormozgan University of Medical Sciences (ethical code: IR.HUMS.REC.1402.395). The study was conducted in accordance with the ethical principles of the Declaration of Helsinki. Written informed consent was obtained from all participants prior to their inclusion in the study. All data were collected and analyzed anonymously to ensure participant confidentiality.

